# Urinary Volatile Organic Compound Metabolites are Associated with High Blood Pressure Among Non-smoking Participants in the National Health and Nutrition Examination Survey (2011-2018)

**DOI:** 10.1101/2024.07.18.24310671

**Authors:** Katlyn E. McGraw, Arce Domingo-Relloso, Daniel W. Riggs, Danielle N. Medgyesi, Raghavee Neupane, Jeanette A. Stingone, Tiffany R. Sanchez

**Affiliations:** Columbia University Mailman School of Public Health Department of Environmental Health Science, 722 W 168th St, New York, NY 10032; Christina Lee Browne Envirome Institute University of Louisville, 322 W Muhammad Ali Blvd, Louisville, KY 40202; Dr. Kiran C. Patel College of Osteopathic Medicine Nova Southeastern University, 3200 S University Dr, Fort Lauderdale, FL 33328; Columbia University Mailman School of Public Health Department of Epidemiology, 722 W 168th St, New York, NY 10032

**Keywords:** Volatile organic compounds, blood pressure, mixtures, acrolein, butadiene

## Abstract

**Background:** Volatile organic compounds (VOCs) are ubiquitous environmental pollutants. Exposure to VOCs is associated with cardiovascular disease (CVD) risk factors, including elevated blood pressure (BP) in susceptible populations. However, research in the general population, particularly among non-smoking adults, is limited. We hypothesized that higher VOC exposure is associated with higher BP and hypertension, among non-smokers.

**Methods:** We included four cycles of data (2011-2018) of non-smoking adults (n=4,430) from the National Health and Nutrition Examination Survey (NHANES). Urinary VOC metabolites were measured by ultra-performance liquid chromatography–mass spectrometry, adjusted for urine dilution, and log-transformed. We estimated mean differences in BP using linear models and prevalence ratio of stage 2 hypertension using modified Poisson models with robust standard errors. Models were adjusted for age, sex, race and ethnicity, education, body mass index, estimated glomerular filtration rate and NHANES cycle.

**Results:** Participants were 54% female, with a median age of 48 years, 32.3% had hypertension, and 7.9% had diabetes. The mean differences (95% CI) in systolic BP were 1.61 (0.07, 3.15) and 2.46 (1.01, 3.92) mmHg when comparing the highest to lowest quartile of urinary acrolein (CEMA) and 1,3-butadiene (DHBMA) metabolites. The prevalence ratios (PR) for hypertension were 1.06 (1.02, 1.09) and 1.05 (1.01, 1.09) when comparing the highest to lowest quartiles of urinary acrolein (CEMA) and 1,3-butadiene (DHBMA), respectively.

**Conclusions:** Exposure to VOCs may be relevant yet understudied environmental contributors to CVD risk in the non-smoking, US population.

## INTRODUCTION

Volatile organic compounds (VOCs) are an under regulated class of chemicals characterized by low molecular weight and high vapor pressure, properties which lead to rapid vaporization in ambient air.^1–4^ Ambient VOCs are critical in the formation of harmful secondary air pollutants such as ozone and PM_2.5_.^5, 6^ However, VOCs emitted directly from sources are just as harmful and include tobacco smoke, vehicle exhaust, industrial solvents, manufacture of consumer goods, and oil and gas refining.^7, 8^ Indoor, VOC levels can be up to ten times higher than ambient VOC levels.^8^ Additionally, some VOCs are produced endogenously through processes like lipid peroxidation, glycation, and amino acid oxidation.^9–11^ Therefore, VOCs are ubiquitous environmental exposures.

Urinary biomarkers of VOC exposure provide proximal, personal-level assessments of all routes of exposure to VOCs, including ambient, indoor, and endogenous sources. VOC metabolites from human biomonitoring have been associated with physician-diagnosed cardiovascular disease (CVD),^12^ metabolic syndrome,^13^ CVD risk factors,^14–17^ and ischemic heart disease mortality,^18^ particularly for acrolein, benzene, 1,3-butadiene, crotonaldehyde, and styrene. However, these studies identified associations in small, geographically homogenous cohorts, and in CVD-susceptible populations, including individuals who smoke. The relation between VOC biomarkers and blood pressure among non-smoking adults has not yet been elucidated in a large, representative sample of the U.S. population, such as the National Health and Nutrition Examination Survey (NHANES).

In this study, we evaluated the association between participants’ VOC metabolite levels in urine with measured blood pressure (BP) levels and hypertension defined as systolic BP ≥140 mmHg, diastolic BP ≥90 mmHg, or self-reported use of BP medications^19^ using data from a representative sample of the non-smoking U.S. adult population participating in NHANES from 2011-2018. We hypothesized that exposure to VOCs, specifically urinary metabolites of acrolein, benzene, 1,3-butadiene, crotonaldehyde, and styrene, would be associated with higher BP levels and prevalence of hypertension. Additionally, we conducted an exploratory assessment of all urinary VOC metabolites measured in NHANES and mixtures of urinary VOC metabolites.

## METHODS

### Study Population

Led by the National Center for Health Statistics (NCHS) at the Centers for Disease Control and Prevention, NHANES is a series of continuous surveys designed to assess the health and nutritional status of adults and children in the U.S. NHANES is designed as a multiyear, stratified, clustered four-stage sample of noninstitutionalized civilians with fixed sample-size targets for sampling domains defined by age, sex, race and ethnicity, and socioeconomic status, with data released in 2-year cycles. Participants gave informed consent of the survey process and their rights as a participant, and the survey was approved by the NCHS Review Board.^20^ Questionnaires were administered in-home followed by standardized health examinations in specially equipped mobile examination centers. Deidentified and detailed health datasets are publicly available on the NHANES website.^21^ We combined NHANES data from four cycles (2011-2018) to create a larger and more geographically diverse sample, including all cycles with available data on BP and urinary VOC metabolites.

### Exclusion Criteria

Of the 39,156 NHANES participants from 2011-2018 combined, 23,825 were 18 years of age and older. We focused on adults because blood pressure varies in childhood and adolescence. There were 7,626 participants with measured urinary VOC metabolites and BP examination data because 1/3 subsample at each cycle are analyzed for environmental chemicals. Because tobacco smoke is a primary source of VOC exposure, self-reported cigarette smoking, and serum cotinine data were used to determine smoking status. We excluded 1,405 individuals who reported currently smoking cigarettes, an additional 451 individuals with serum cotinine levels >10 µg/L, reflecting recent nicotine exposure, and 480 missing self-reported cigarette smoking status or cotinine data, for a final sample of 5,290 non-smoking participants. We further excluded an additional 860 participants with missing data, including 492 missing priority urinary VOCs, 172 missing systolic blood pressure (SBP), 11 missing diastolic blood pressure (DBP), 81 missing urine creatinine, 42 missing BMI, 35 missing diabetes status, 21 missing estimated glomerular filtration rate (eGFR), 3 missing triglycerides (TG), 2 missing high density lipoproteins (HDL), and 1 missing education, for a total of 4,430 participants included in our analysis (Figure S1).

### Urinary VOC Metabolites

Urine specimens were collected at mobile examination centers, frozen at -20°C, and shipped to the Division of Environmental Health Laboratory Sciences, National Center for Environmental Health for analysis.^22^ Ultra-performance liquid chromatography coupled with electrospray tandem mass spectrometry was used to measure 35 VOC metabolites of 20 parent compounds in urine in nanograms per milliliter (ng/mL).^23^

We report results for 5 hypothesized, priority parent VOCs and their 9 associated urinary metabolites: acrolein (CEMA – N-Acetyl-S-(2-carboxyethyl)-L-cysteine and 3HPMA – N-Acetyl-S-(3-hydroxypropyl)-L-cysteine), benzene (MU – t,t-Muconic Acid and PMA – N-Acetyl-S-(phenyl)-L-cysteine), 1,3-butadiene (DHBMA – N-Acetyl-S-(3,4-dihydroxybutyl)-L-cysteine and MHBMA3 – N-Acetyl-S-(4-hydroxy-2-butenyl)-L-cysteine), crotonaldehyde (HPMMA – N-Acetyl-S-(3-hydroxypropyl-1-methyl)-L-cysteine), and styrene (PGA – N-Acetyl-S-(phenyl)-L-cysteine and MA – Mandelic acid). PGA and MA are metabolites of ethylbenzene and styrene. See Table S1 for all parent compounds, long and short metabolite names, NHANES cycle availability, and limits of detection (LOD). Values below the LOD were imputed with the LOD/√2. Although the benzene metabolite, MU, was only available for one cycle (2017-2018, n=1,046), we included MU in primary analyses for comparison to the other benzene metabolite available at the other three cycles, PMA (2011-2016, n=3,207). To correct for urine dilution, we divided all urinary VOC metabolites by urine creatinine and reported levels as nanogram of VOC metabolite per milligrams of creatinine (ng/mg).

### Blood Pressure and Hypertension

BP measurements were collected at mobile examination centers using a sphygmomanometer and appropriately sized cuff for the participant’s arm circumference. Three BP readings were collected after five minutes of seated rest. A fourth measurement was collected if a measurement was interrupted or incomplete. All recorded measurements of SBP and DBP were averaged and used as continuous outcome variables.

For hypertension, our primary endpoint was defined by having either a mean SBP ≥140 mmHg, a mean DBP ≥90 mmHg, or self-reported use of BP medications , also known as stage 2 hypertension.^19^ We also conducted sensitivity analyses using two alternate definitions: 1) stage 1 hypertension, where participants were hypertensive if mean SBP ≥130 mmHg, mean DBP ≥80 mmHg, or were taking BP medications, and 2) if individuals self-reported physician-diagnosed hypertension.

### Covariates

Age, sex, race and ethnicity, education, and household income were acquired from self-reported questionnaires. Race and ethnicity were classified as Mexican American, Other Hispanic, Non-Hispanic White, Non-Hispanic Black, Non-Hispanic Asian, and Other Races, including multiple races. We reclassified education as less than a high school education, a high school education or GED equivalent, some college, or bachelor’s degree or more. Household income categories were reduced to $0-$24,999, $25,000-$54,999, $55,000-$74,999, and ≥$75,000. Body mass index (BMI, kg/m^2^) was collected during examination at mobile examination center. Estimated glomerular filtration rate (eGFR) was calculated using the Chronic Kidney Disease Epidemiology Collaboration equation, ^24^ which estimates GFR for age, sex, and serum creatinine in µmol/L. The eGFR can influence metabolite excretion in urine and was therefore used for adjustment in our models. Total cholesterol and high-density lipoprotein (HDL) cholesterol were measured in blood collected at mobile examination centers. Diabetes status was determined based on self-reported responses that the participant was ever told by a doctor they had diabetes or were taking medications to lower blood sugar. Prevalent CVD status was determined based on self-reported responses that the participant was ever told by a doctor they had congestive heart failure, coronary heart disease, angina or angina pectoris, heart attack, or stroke.

### Statistics

Combined NHANES survey cycles and weights produce estimates representative of the U.S. civilian noninstitutionalized population at the midpoint of the combined survey period. We constructed new sample weights for the combined cycles (4 cycles, 2011-2018) by multiplying 2-year subsample weights for environmental chemicals by 1/4 (for 4 NHANES cycles) as described.^25^

To characterize our study population, we stratified by hypertension status and continuous and categorical variables of interest using t-tests and χ² tests. We used linear regression models for normally distributed BP and modified Poisson regression with robust standard errors for dichotomous hypertension outcomes to estimate prevalence ratios of hypertension.^26^ Urine VOC metabolite levels were modeled as: (1) quartiles (to compare each of the highest three quartiles to the lowest quartile), (2) per interquartile range (IQR) on log-transformed levels (to compare the 75th to the 25th percentile), and (3) log-transformed levels with restricted quadratic splines (to evaluate the flexible dose-response relationship). Because NHANES represents a younger, more general population, we chose to adjust for sociodemographic factors age, sex, race and ethnicity, education, eGFR, and NHANES cycle year and BMI in our main model, Model 1, as to not adjust away potential mediating CVD risk factors based on literature reviews for BP and hypertension.^27, 28^ In Model 2, we further adjust for traditional CVD risk factors including total cholesterol, HDL, triglycerides, diabetes status, and BP medications. We present 9 priority urinary VOC metabolites of parent VOCs acrolein, benzene, 1,3-butadiene, crotonaldehyde, and styrene, from Model 1, in the main text. Model estimates are population weighted mean differences in BP or population weighted prevalence ratio of hypertension with 95% confidence intervals (CI). All central tendency estimates, proportions and effect estimates are population weighted.

### Secondary analyses

In secondary analyses, we examined the other 12 VOC metabolites measured in NHANES. We used Wald tests and conducted subgroup analysis to assess effect modification by subgroups of age, sex, and race and ethnicity.

To address potential co-pollutant exposure and collinearity, we used hierarchical Bayesian Kernel Machine Regression (BKMR). Because BKMR cannot account for complex survey weighting, we consider these exploratory analyses. BKMR is a kernel-regression based machine learning method that characterizes the exposure response function of multiple predictors on a health outcome while other predictors are fixed to a specific percentile.^29, 30^ The method also allows us to examine statistical interactions between VOC metabolites within the mixture and joint associations between the whole mixture and health outcomes. The hierarchical BKMR uses hierarchical variable selection to create posterior inclusion probabilities (PIPs) that quantify the relative importance by selecting 1) at the group level and 2) by selecting exposures of each group.^29^ All metabolites with the same parent compound were grouped in hierarchical analysis except for MHBMA3 and HPMMA which were more highly correlated than DHBMA and MHBMA3 (Figure S2). We excluded the benzene metabolites, MU and PMA, due to missing cycles. BKMRs were run with 10,000 iterations and the tuning parameter r.jump value of 0.1 for continuous SBP and DBP and dichotomous hypertension to achieve higher acceptance rates.

### Sensitivity Analyses

We conducted several sensitivity analyses. Because a number of participants reported use of BP lowering medications, we assessed differences between measured and underlying BP.^31^ For individuals self-reporting use of BP lowering medications we imputed underlying BP values by adding a 10 mm Hg or 5 mm Hg constant to measured SBP or DBP, respectively.^31^ Additionally, we assessed differences between Stage 1, Stage 2, and physician-diagnosed hypertension.

Data analysis was performed in R (version 3.1.3)^32^ using the nhanesA^33^, tidyverse^34^ and survey package^35^ to account for the complex survey design and sampling weights.

## RESULTS

The characteristics of NHANES participants by hypertension status are shown in Table 1. Participants were 54% female, with a median age of 48 years, 32.3% had hypertension, and 17.9% had diabetes mellitus. Participants with hypertension were older and more likely to be non-Hispanic White or non-Hispanic Black, had higher BMI, lower eGFR, lower HDL cholesterol, higher triglycerides, higher total cholesterol, and were more likely to have self-reported physician diagnosed CVD and diabetes mellitus. Additionally, participants with hypertension had higher median levels of all priority urinary VOC metabolites compared to participants without hypertension (Table 2 and Table S2), except for the PMA benzene metabolite.

**Table 1.** Participant characteristics across categories of hypertension, restricted to non-smoking participants with available blood pressure examination data and urinary VOCs, n=4,430 NHANES 2011-2018 participants. Continuous variables are reported as median [interquartile range, IQR] and categorical variables as number (%). Percentages are population weighted.

|  | Overall | No Hypertension | Hypertension |
| --- | --- | --- | --- |
| n | 4,430 | 2,819 | 1,611 |
| Age (years) | 48 [33, 62] | 40 [28, 54] | 62 [54, 71] |
| Sex |  |  |  |
| Male | 2,044 (46.0) | 1,259 (45.3) | 785 (47.5) |
| Female | 2,386 (54.0) | 1,560 (54.7) | 826 (52.5) |
| Race/Ethnicity |  |  |  |
| Mexican American | 681 (9.6) | 494 (11.3) | 187 (6.1) |
| Other Hispanic | 533 (7.3) | 369 (8.4) | 164 (5.0) |
| Non-Hispanic White | 1,540 (64.8) | 943 (62.8) | 597 (69.0) |
| Non-Hispanic Black | 868 (9.7) | 449 (8.3) | 419 (12.6) |
| Non-Hispanic Asian | 679 (6.3) | 478 (6.8) | 201 (5.2) |
| Other Race, incl Multi | 129 (2.4) | 86 (2.5) | 43 (2.2) |
| Education |  |  |  |
| Less than HS Diploma | 884 (12.2) | 500 (11.3) | 384 (14.0) |
| High School Grad/GED | 926 (20.7) | 553 (19.3) | 373 (23.6) |
| Some College | 1,329 (31.1) | 841 (30.1) | 488 (33.3) |
| College Degree or More | 1,291 (36.0) | 925 (39.4) | 366 (28.9) |
| Income |  |  |  |
| \$0-\$24,999 | 1,002 (15.9) | 563 (14.6) | 439 (18.7) |
| \$25,000-\$54,999 | 1,308 (29.1) | 828 (29.4) | 480 (28.5) |
| \$55,000-\$74,999 | 490 (13.1) | 335 (13.4) | 155 (12.4) |
| ≥\$75,000 | 1,352 (41.9) | 913 (42.7) | 439 (40.4) |
| BMI (kg/m<sup>2</sup>) | 28.3 [24.5, 33.0] | 27.2 [23.7, 31.7] | 30.7 [26.6, 35.8] |
| Serum Cotinine (ng/mL) | 0.02 [0.01, 0.05] | 0.02 [0.01, 0.05] | 0.02 [0.01, 0.04] |
| eGFR (mL/min/1.73 m<sup>2</sup>) | 97.6 [82.0, 111.9] | 103.3 [88.4, 116.7] | 86.2 [68.9, 98.9] |
| HDL (mg/dL) | 53.0 [43.0, 63.0] | 53.3 [44.0, 64.0] | 50.0 [41.0, 61.0] |
| Triglycerides (mg/dL) | 115.0 [77.0, 176.0] | 104.0 [72.0, 165.0] | 134.0 [94.0, 206.0] |
| Total Cholesterol (mg/dL) | 186.0 [161.0, 214.0] | 185.0 [160.0, 213.0] | 191.5 [163.0, 218.0] |
| SBP (mmHg) | 120.0 [110.7, 132.0] | 114.7 [107.3, 123.3] | 136.0 [124.0, 147.3] |
| DBP (mmHg) | 71.3 [64.7, 78.0] | 70.0 [64.0, 76.0] | 75.3 [66.7, 84.0] |
| Underlying SBP (mmHg)<sup>a</sup> | 122.0 [111.3, 135.3] | 114.7 [107.3, 123.3] | 142.7 [133.3, 152.7] |
| Underlying DBP (mmHg)<sup>b</sup> | 72.3 [65.3, 79.3] | 70.0 [64.0, 76.0] | 79.0 [71.0, 87.0] |
| Hypertension Stage 1<sup>c</sup> | 2,201 (46.9) | 590 (21.5) | 1,611 (100.0) |
| Hypertension MDDX<sup>d</sup> | 1,542 (31.2) | 216 (7.0) | 1,326 (81.9) |
| CVD<sup>e</sup> | 382 (7.6) | 100 (3.8) | 282 (15.3) |
| Diabetes Mellitus<sup>f</sup> | 439 (7.9) | 126 (3.1) | 313 (17.9) |
| BP Medications | 1,213 (24.1) | 0 (0.0) | 1,213 (74.5) |

<sup>a</sup> Underlying Systolic Blood Pressure: a 10 mmHg constant was added to SBP measurement of NHANES participants who self-report taking BP medications <sup>b</sup>
Underlying Diastolic Blood Pressure: a 5 mmHg constant was added to SBP measurement of NHANES participants who self-report taking BP medications
<sup>c</sup> Hypertension Stage 1 is defined as mean SBP ≥ 130 mmHg, DBP ≥ 80 mm Hg, or taking BP medications
<sup>d</sup> Hypertension MDDX is defined as physician-diagnosed hypertension
<sup>e</sup> CVD is defined if a physician has ever told you have congestive heart failure, coronary heart disease, angina, heart attack, or stroke
<sup>f</sup> Diabetes Mellitus is defined if a physician ever told you have diabetes mellitus or self-report of taking diabetes medications

**Table 2.** Median and interquartile ranges of VOC metabolite levels (ng/mg creatinine) across categories of hypertension, restricted to participants with available blood pressure examination data and urinary VOCs, among 4,430 NHANES 2011-2018 participants. MU was measured in n=1,046 NHANES 2017-2018 participants and PMA was measured in n=3,207 NHANES 2011-2016 participants.

| Parent | Short<sup>a</sup> | Overall | No Hypertension | Hypertension |
| --- | --- | --- | --- | --- |
|  |  | 4,430 | 2,819 | 1,611 |
| Acrolein | CEMA | 82.3 [54.5, 128.2] | 76.2 [50.4, 116.7] | 96.7 [64.5, 150.4] |
| Acrolein | 3HPMA | 193.7 [130.4, 297.6] | 189.6 [127.4, 290.0] | 202.8 [137.6, 310.8] |
| Benzene | MU<sup>b</sup> | 44.8 [26.8, 93.0] | 43.3 [24.5, 93.0] | 47.7 [29.0, 93.1] |
| Benzene | PMA<sup>c</sup> | 0.7 [0.5, 1.2] | 0.8 [0.4, 1.3] | 0.7 [0.5, 1.1] |
| 1,3-Butadiene | DHBMA | 300.0 [236.3, 385.1] | 288.6 [227.5, 363.5] | 340.1 [264.1, 429.8] |
| 1,3-Butadiene | MHBMA3 | 3.9 [2.6, 6.0] | 3.7 [2.5, 5.9] | 4.2 [2.9, 6.2] |
| Crotonaldehyde | HPMMA | 185.5 [143.0, 264.6] | 173.6 [136.9, 250.0] | 212.7 [159.3, 298.8] |
| Styrene | PGA | 197.8 [148.5, 260.4] | 190.6 [142.1, 251.0] | 214.1 [158.4, 281.3] |
| Styrene | MA | 122.4 [89.5, 164.4] | 120.2 [88.9, 159.2] | 128.4 [91.9, 172.9] |

<sup>a</sup> Metabolite long names: CEMA, N-Acetyl-S-(2-carboxyethyl)-L-cysteine; 3HPMA, N-Acetyl-S-(3-hydroxypropyl)-L-cysteine; PMA, N-Acetyl-S-(phenyl)-L-cysteine; MU, *t,t*-Muconic Acid; DHBMA, N-Acetyl-S-(3,4-dihydroxybutyl)-L-cysteine; MHBMA3, N-Acetyl-S-(4-hydroxy-2-butenyl)-L-cysteine; HPMMA, N-Acetyl-S-(3-hydroxypropyl-1-methyl)-L-cysteine; PGA, Phenylglyoxylic acid; MA, Mandelic acid
<sup>b</sup> MU was measured in n=1,046 NHANES 2017-2018 participants, n=604 did not have hypertension and n=442 did have hypertension
<sup>c</sup> PMA was measured in n=3,207 NHANES 2011-2016 participants, n=2,104 did not have hypertension and n=1,103 did have hypertension

Results for priority VOC metabolites and BP outcomes are shown in Table 3. For acrolein, the urinary metabolite CEMA was associated with higher SBP and a higher prevalence of hypertension. When comparing the highest to lowest quartile, the mean difference (MD) (95%CI) was 1.61 (0.07, 3.15) mmHg for SBP, and the prevalence ratio (PR) (95%CI) of hypertension was 1.06 (1.02, 1.09). Estimates for SBP and hypertension were also statistically significant per log IQR-change and, although attenuated, generally remained significant after further adjustment for CVD risk factors. In the restricted quadratic spline models, there were clear positive dose-response relationships with SBP and PR of hypertension observed for CEMA above 34.20 ng/mg creatinine (Figures 1 and 2). For the other acrolein metabolite, 3HPMA, results were positive but not significant for SBP (MD [95%CI]: 1.28 [-0.33, 2.90] mmHg); however, the association with hypertension was statistically significant (PR [95%CI]: 1.05 [1.01, 1.09]).

**Figure 1.**
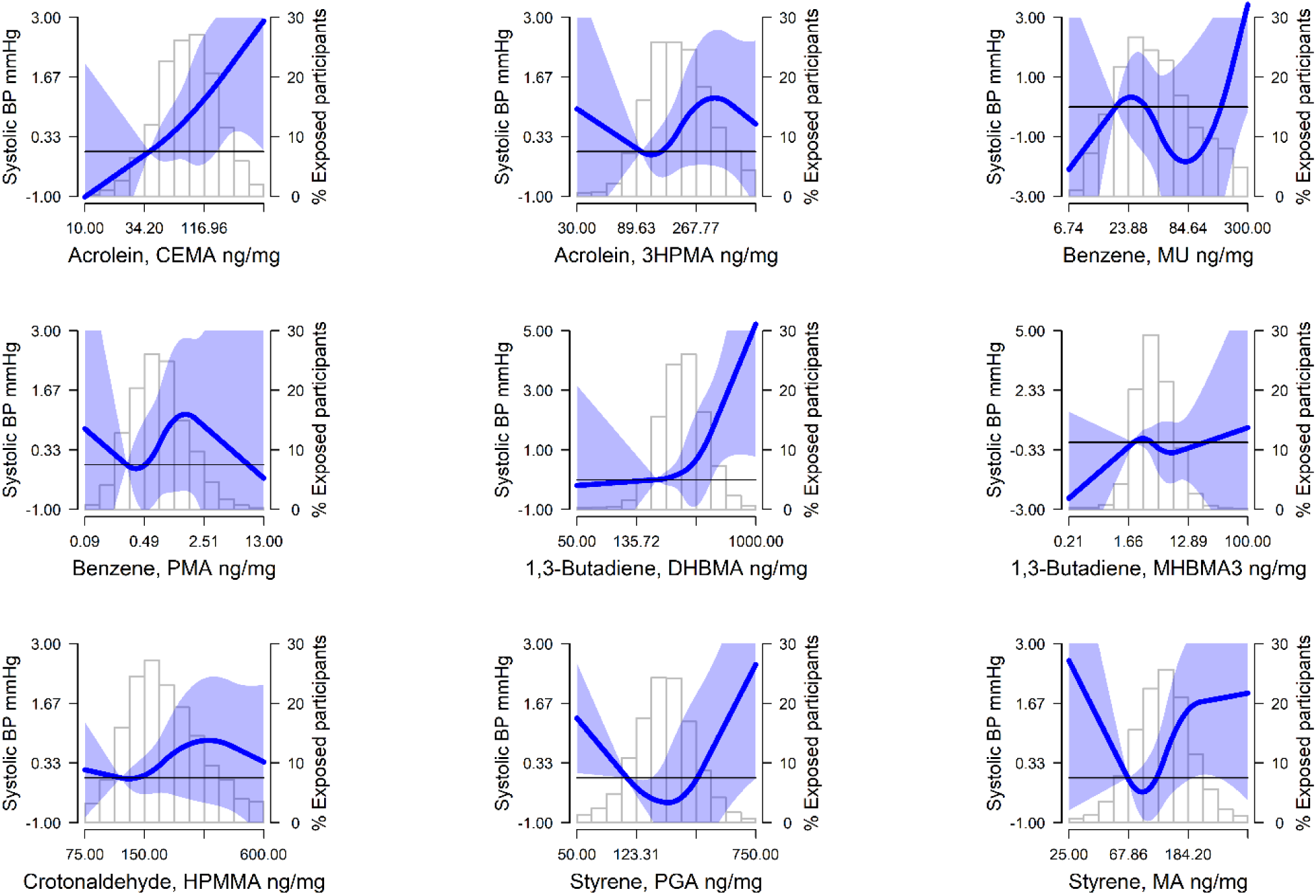
Mean differences (MD) (95% confidence interval) of systolic blood pressure (SBP) by urinary VOC levels (ng/mg creatinine) modeled as restricted quadratic splines for priority VOC metabolites among nonsmoking NHANES 2011-2018 participants (n=4,430). Lines (shaded areas) represent the MD (95%CI) of SBP and RR (95% CI) of hypertension by VOC metabolites modeled as restricted quadratic splines for log transformed VOC metabolite distributions with knots at 10th, 50th, and 90th percentiles. The reference value was set at the 10th percentile. Models were adjusted for age, sex, race and ethnicity, education, BMI, eGFR, and NHANES cycle year. The histograms in the background represent the distribution of each VOC metabolite (ng/mg creatinine). See Figure S2 for DBP.

**Figure 2.**
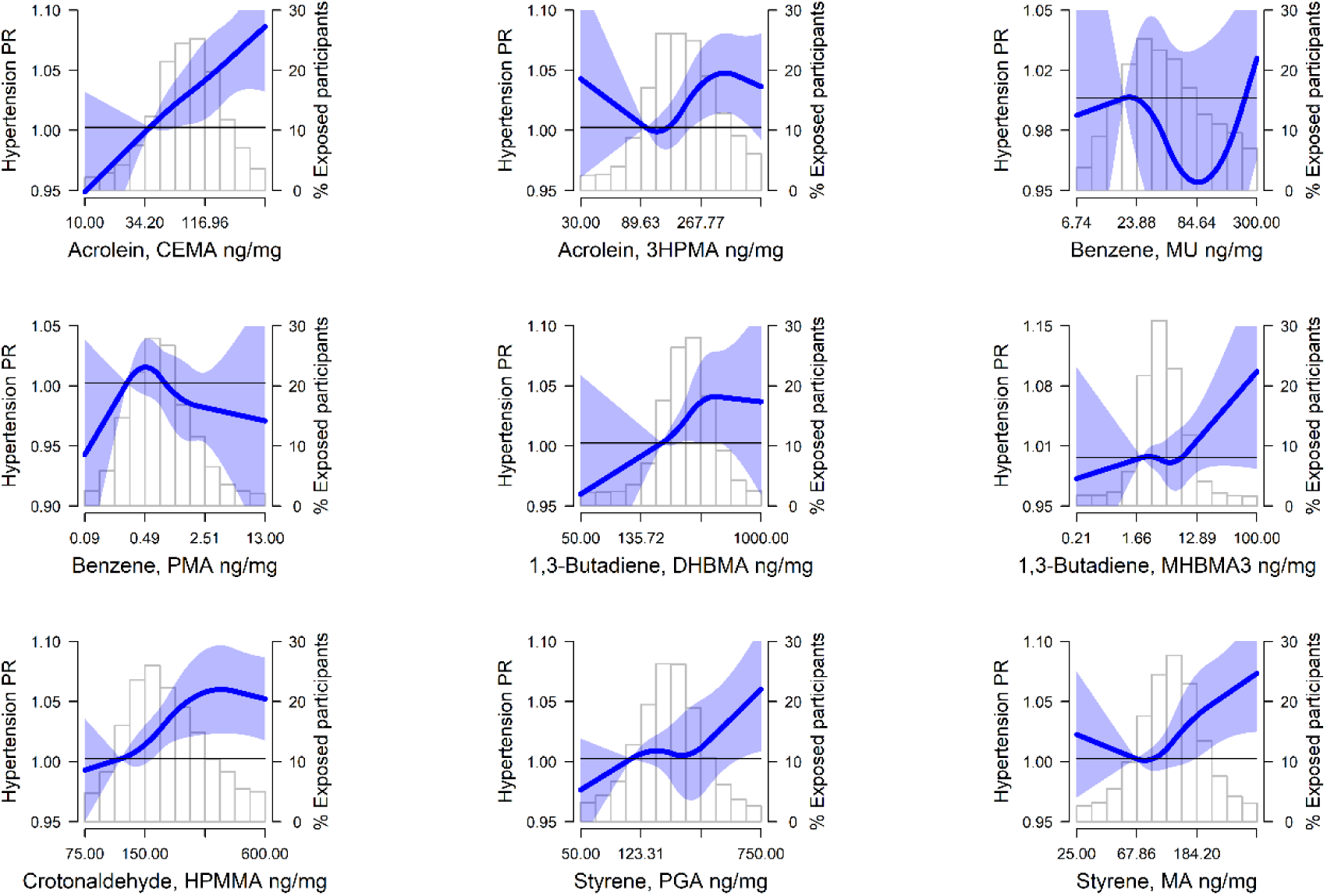
Prevalence ratios (PR) (95% confidence interval, CI) of hypertension by urinary VOC levels (ng/mg creatinine) modeled as restricted quadratic splines among nonsmoking NHANES 2011-2018 participants (n=4,430). Lines (shaded areas) represent the PR (95%CI) of hypertension by VOC metabolites modeled as restricted quadratic splines for log transformed VOC metabolite distributions with knots at 10th, 50th, and 90th percentiles. The reference value was set at the 10th percentile. Models were adjusted for age, sex, race and ethnicity, education, BMI, eGFR, and NHANES cycle year. The histograms in the background represent the distribution of each VOC metabolite (ng/mg creatinine). See Figure S2 for DBP.

**Table 3.**
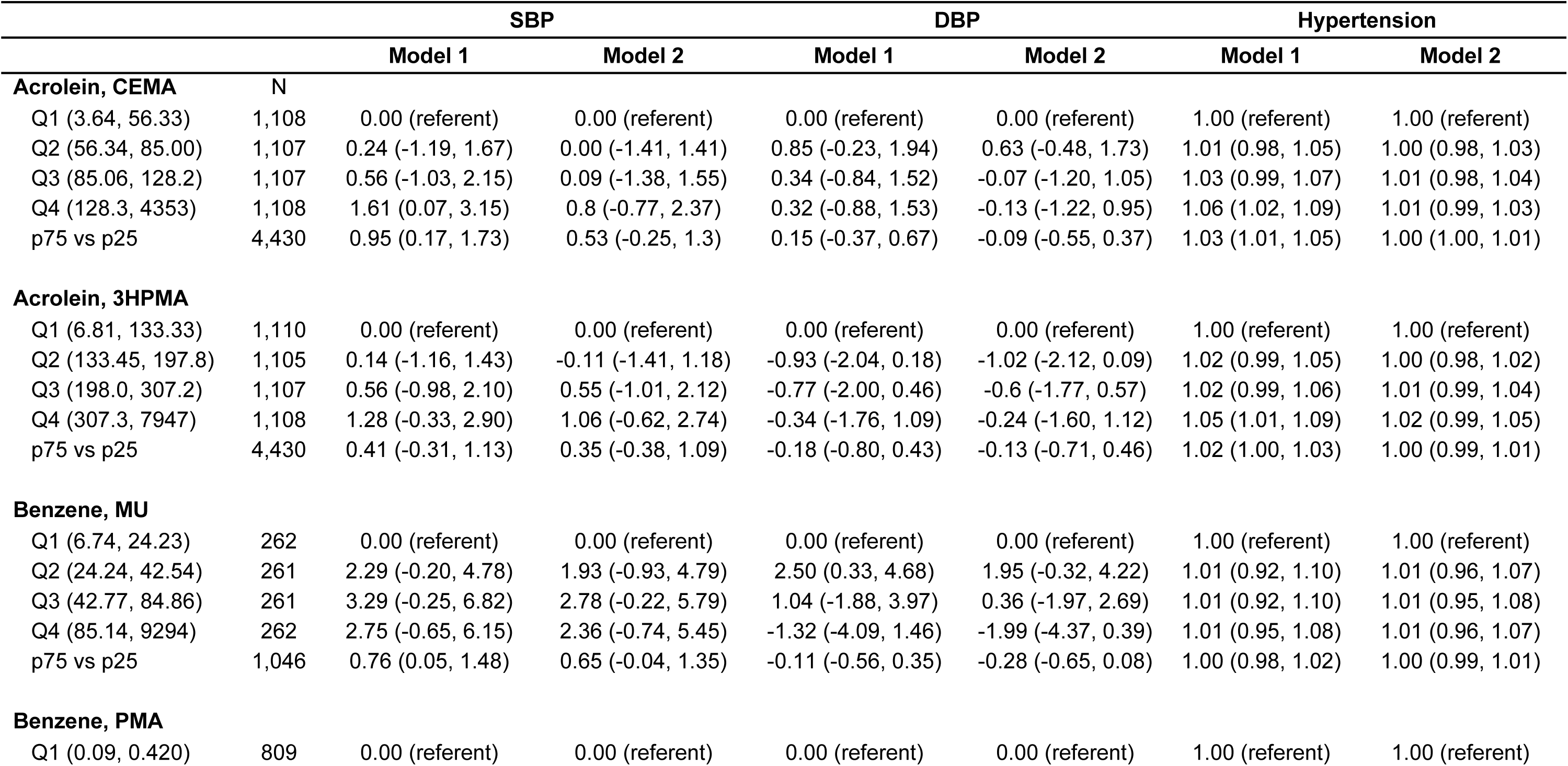
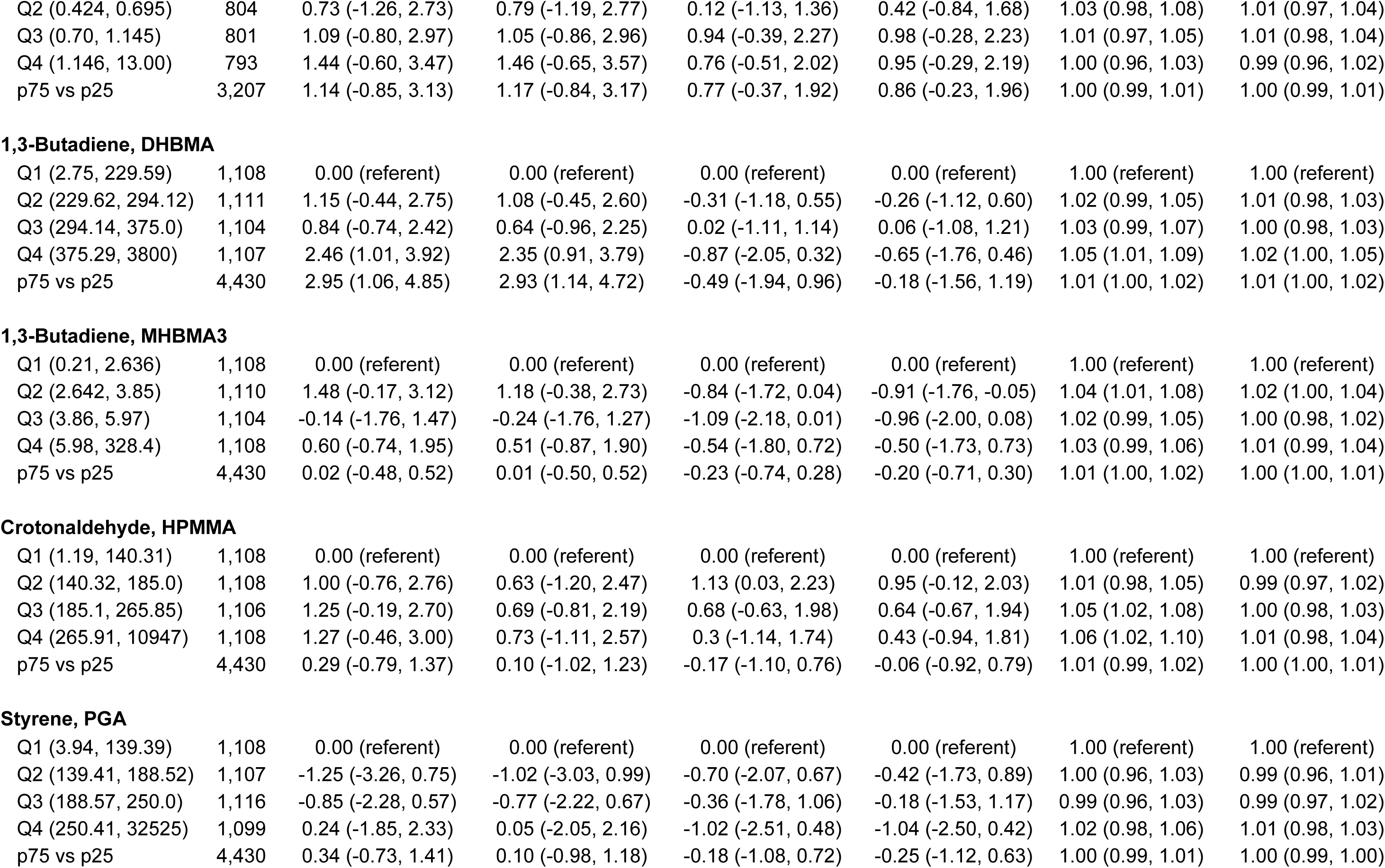
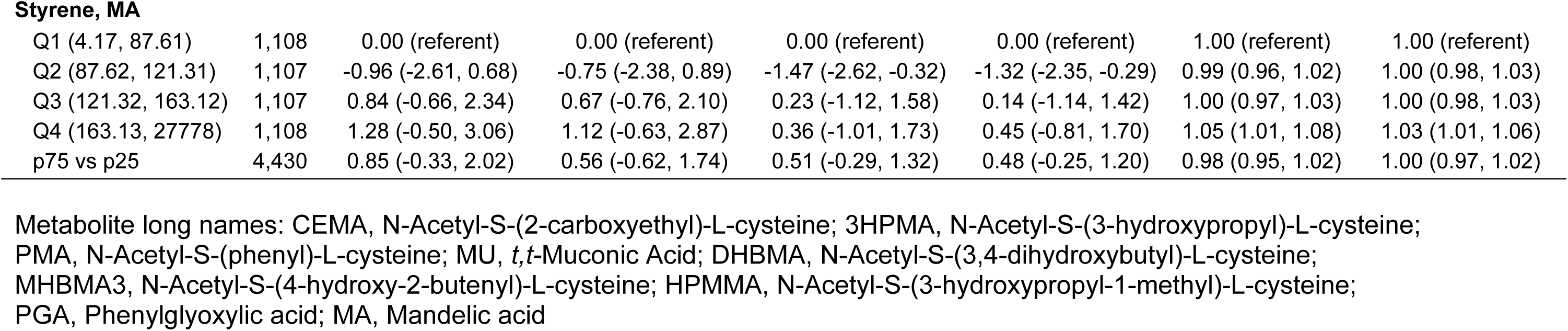
Mean differences (95% confidence interval) of systolic blood pressure (SBP), diastolic blood pressure (DBP), and prevalence ratios – PR (95% confidence interval) of hypertension by levels (ng/mg creatinine) of urinary VOC metabolites, restricted to participants with available blood pressure examination data and urinary VOCs, n=4,430 NHANES 2011-2018 participants. MU was measured in n=1,046 NHANES 2017-2018 participants and PMA was measured in n=3,207 NHANES 2011-2016 participants. Model 1 was adjusted for age, sex, race and ethnicity, education, BMI, eGFR, and NHANES cycle year. Model 2 was additionally adjusted for HDL-cholesterol, triglycerides, total cholesterol, diabetes status, and antihypertensive medication.

|  |  | SBP |  | DBP |  | Hypertension |  |
| --- | --- | --- | --- | --- | --- | --- | --- |
|  |  | Model 1 | Model 2 | Model 1 | Model 2 | Model 1 | Model 2 |
| **Acrolein, CEMA** |  |  |  |  |  |  |  |
|  | N |  |  |  |  |  |  |
| Q1 (3.64, 56.33) | 1,108 | 0.00 (referent) | 0.00 (referent) | 0.00 (referent) | 0.00 (referent) | 1.00 (referent) | 1.00 (referent) |
| Q2 (56.34, 85.00) | 1,107 | 0.24 (-1.19, 1.67) | 0.00 (-1.41, 1.41) | 0.85 (-0.23, 1.94) | 0.63 (-0.48, 1.73) | 1.01 (0.98, 1.05) | 1.00 (0.98, 1.03) |
| Q3 (85.06, 128.2) | 1,107 | 0.56 (-1.03, 2.15) | 0.09 (-1.38, 1.55) | 0.34 (-0.84, 1.52) | -0.07 (-1.20, 1.05) | 1.03 (0.99, 1.07) | 1.01 (0.98, 1.04) |
| Q4 (128.3, 4353) | 1,108 | 1.61 (0.07, 3.15) | 0.8 (-0.77, 2.37) | 0.32 (-0.88, 1.53) | -0.13 (-1.22, 0.95) | 1.06 (1.02, 1.09) | 1.01 (0.99, 1.03) |
| p75 vs p25 | 4,430 | 0.95 (0.17, 1.73) | 0.53 (-0.25, 1.3) | 0.15 (-0.37, 0.67) | -0.09 (-0.55, 0.37) | 1.03 (1.01, 1.05) | 1.00 (1.00, 1.01) |
| **Acrolein, 3HPMA** |  |  |  |  |  |  |  |
| Q1 (6.81, 133.33) | 1,110 | 0.00 (referent) | 0.00 (referent) | 0.00 (referent) | 0.00 (referent) | 1.00 (referent) | 1.00 (referent) |
| Q2 (133.45, 197.8) | 1,105 | 0.14 (-1.16, 1.43) | -0.11 (-1.41, 1.18) | -0.93 (-2.04, 0.18) | -1.02 (-2.12, 0.09) | 1.02 (0.99, 1.05) | 1.00 (0.98, 1.02) |
| Q3 (198.0, 307.2) | 1,107 | 0.56 (-0.98, 2.10) | 0.55 (-1.01, 2.12) | -0.77 (-2.00, 0.46) | -0.6 (-1.77, 0.57) | 1.02 (0.99, 1.06) | 1.01 (0.99, 1.04) |
| Q4 (307.3, 7947) | 1,108 | 1.28 (-0.33, 2.90) | 1.06 (-0.62, 2.74) | -0.34 (-1.76, 1.09) | -0.24 (-1.60, 1.12) | 1.05 (1.01, 1.09) | 1.02 (0.99, 1.05) |
| p75 vs p25 | 4,430 | 0.41 (-0.31, 1.13) | 0.35 (-0.38, 1.09) | -0.18 (-0.80, 0.43) | -0.13 (-0.71, 0.46) | 1.02 (1.00, 1.03) | 1.00 (0.99, 1.01) |
| **Benzene, MU** |  |  |  |  |  |  |  |
| Q1 (6.74, 24.23) | 262 | 0.00 (referent) | 0.00 (referent) | 0.00 (referent) | 0.00 (referent) | 1.00 (referent) | 1.00 (referent) |
| Q2 (24.24, 42.54) | 261 | 2.29 (-0.20, 4.78) | 1.93 (-0.93, 4.79) | 2.50 (0.33, 4.68) | 1.95 (-0.32, 4.22) | 1.01 (0.92, 1.10) | 1.01 (0.96, 1.07) |
| Q3 (42.77, 84.86) | 261 | 3.29 (-0.25, 6.82) | 2.78 (-0.22, 5.79) | 1.04 (-1.88, 3.97) | 0.36 (-1.97, 2.69) | 1.01 (0.92, 1.10) | 1.01 (0.95, 1.08) |
| Q4 (85.14, 9294) | 262 | 2.75 (-0.65, 6.15) | 2.36 (-0.74, 5.45) | -1.32 (-4.09, 1.46) | -1.99 (-4.37, 0.39) | 1.01 (0.95, 1.08) | 1.01 (0.96, 1.07) |
| p75 vs p25 | 1,046 | 0.76 (0.05, 1.48) | 0.65 (-0.04, 1.35) | -0.11 (-0.56, 0.35) | -0.28 (-0.65, 0.08) | 1.00 (0.98, 1.02) | 1.00 (0.99, 1.01) |
| **Benzene, PMA** |  |  |  |  |  |  |  |
| Q1 (0.09, 0.420) | 809 | 0.00 (referent) | 0.00 (referent) | 0.00 (referent) | 0.00 (referent) | 1.00 (referent) | 1.00 (referent) |
| Q2 (0.424, 0.695) | 804 | 0.73 (-1.26, 2.73) | 0.79 (-1.19, 2.77) | 0.12 (-1.13, 1.36) | 0.42 (-0.84, 1.68) | 1.03 (0.98, 1.08) | 1.01 (0.97, 1.04) |
| Q3 (0.70, 1.145) | 801 | 1.09 (-0.80, 2.97) | 1.05 (-0.86, 2.96) | 0.94 (-0.39, 2.27) | 0.98 (-0.28, 2.23) | 1.01 (0.97, 1.05) | 1.01 (0.98, 1.04) |
| Q4 (1.146, 13.00) | 793 | 1.44 (-0.60, 3.47) | 1.46 (-0.65, 3.57) | 0.76 (-0.51, 2.02) | 0.95 (-0.29, 2.19) | 1.00 (0.96, 1.03) | 0.99 (0.96, 1.02) |
| p75 vs p25 | 3,207 | 1.14 (-0.85, 3.13) | 1.17 (-0.84, 3.17) | 0.77 (-0.37, 1.92) | 0.86 (-0.23, 1.96) | 1.00 (0.99, 1.01) | 1.00 (0.99, 1.01) |
| 1,3-Butadiene, DHBMA |  |  |  |  |  |  |  |
| Q1 (2.75, 229.59) | 1,108 | 0.00 (referent) | 0.00 (referent) | 0.00 (referent) | 0.00 (referent) | 1.00 (referent) | 1.00 (referent) |
| Q2 (229.62, 294.12) | 1,111 | 1.15 (-0.44, 2.75) | 1.08 (-0.45, 2.60) | -0.31 (-1.18, 0.55) | -0.26 (-1.12, 0.60) | 1.02 (0.99, 1.05) | 1.01 (0.98, 1.03) |
| Q3 (294.14, 375.0) | 1,104 | 0.84 (-0.74, 2.42) | 0.64 (-0.96, 2.25) | 0.02 (-1.11, 1.14) | 0.06 (-1.08, 1.21) | 1.03 (0.99, 1.07) | 1.00 (0.98, 1.03) |
| Q4 (375.29, 3800) | 1,107 | 2.46 (1.01, 3.92) | 2.35 (0.91, 3.79) | -0.87 (-2.05, 0.32) | -0.65 (-1.76, 0.46) | 1.05 (1.01, 1.09) | 1.02 (1.00, 1.05) |
| p75 vs p25 | 4,430 | 2.95 (1.06, 4.85) | 2.93 (1.14, 4.72) | -0.49 (-1.94, 0.96) | -0.18 (-1.56, 1.19) | 1.01 (1.00, 1.02) | 1.01 (1.00, 1.02) |
| 1,3-Butadiene, MHBMA3 |  |  |  |  |  |  |  |
| Q1 (0.21, 2.636) | 1,108 | 0.00 (referent) | 0.00 (referent) | 0.00 (referent) | 0.00 (referent) | 1.00 (referent) | 1.00 (referent) |
| Q2 (2.642, 3.85) | 1,110 | 1.48 (-0.17, 3.12) | 1.18 (-0.38, 2.73) | -0.84 (-1.72, 0.04) | -0.91 (-1.76, -0.05) | 1.04 (1.01, 1.08) | 1.02 (1.00, 1.04) |
| Q3 (3.86, 5.97) | 1,104 | -0.14 (-1.76, 1.47) | -0.24 (-1.76, 1.27) | -1.09 (-2.18, 0.01) | -0.96 (-2.00, 0.08) | 1.02 (0.99, 1.05) | 1.00 (0.98, 1.02) |
| Q4 (5.98, 328.4) | 1,108 | 0.60 (-0.74, 1.95) | 0.51 (-0.87, 1.90) | -0.54 (-1.80, 0.72) | -0.50 (-1.73, 0.73) | 1.03 (0.99, 1.06) | 1.01 (0.99, 1.04) |
| p75 vs p25 | 4,430 | 0.02 (-0.48, 0.52) | 0.01 (-0.50, 0.52) | -0.23 (-0.74, 0.28) | -0.20 (-0.71, 0.30) | 1.01 (1.00, 1.02) | 1.00 (1.00, 1.01) |
| Crotonaldehyde, HPMMA |  |  |  |  |  |  |  |
| Q1 (1.19, 140.31) | 1,108 | 0.00 (referent) | 0.00 (referent) | 0.00 (referent) | 0.00 (referent) | 1.00 (referent) | 1.00 (referent) |
| Q2 (140.32, 185.0) | 1,108 | 1.00 (-0.76, 2.76) | 0.63 (-1.20, 2.47) | 1.13 (0.03, 2.23) | 0.95 (-0.12, 2.03) | 1.01 (0.98, 1.05) | 0.99 (0.97, 1.02) |
| Q3 (185.1, 265.85) | 1,106 | 1.25 (-0.19, 2.70) | 0.69 (-0.81, 2.19) | 0.68 (-0.63, 1.98) | 0.64 (-0.67, 1.94) | 1.05 (1.02, 1.08) | 1.00 (0.98, 1.03) |
| Q4 (265.91, 10947) | 1,108 | 1.27 (-0.46, 3.00) | 0.73 (-1.11, 2.57) | 0.3 (-1.14, 1.74) | 0.43 (-0.94, 1.81) | 1.06 (1.02, 1.10) | 1.01 (0.98, 1.04) |
| p75 vs p25 | 4,430 | 0.29 (-0.79, 1.37) | 0.10 (-1.02, 1.23) | -0.17 (-1.10, 0.76) | -0.06 (-0.92, 0.79) | 1.01 (0.99, 1.02) | 1.00 (1.00, 1.01) |
| Styrene, PGA |  |  |  |  |  |  |  |
| Q1 (3.94, 139.39) | 1,108 | 0.00 (referent) | 0.00 (referent) | 0.00 (referent) | 0.00 (referent) | 1.00 (referent) | 1.00 (referent) |
| Q2 (139.41, 188.52) | 1,107 | -1.25 (-3.26, 0.75) | -1.02 (-3.03, 0.99) | -0.70 (-2.07, 0.67) | -0.42 (-1.73, 0.89) | 1.00 (0.96, 1.03) | 0.99 (0.96, 1.01) |
| Q3 (188.57, 250.0) | 1,116 | -0.85 (-2.28, 0.57) | -0.77 (-2.22, 0.67) | -0.36 (-1.78, 1.06) | -0.18 (-1.53, 1.17) | 0.99 (0.96, 1.03) | 0.99 (0.97, 1.02) |
| Q4 (250.41, 32525) | 1,099 | 0.24 (-1.85, 2.33) | 0.05 (-2.05, 2.16) | -1.02 (-2.51, 0.48) | -1.04 (-2.50, 0.42) | 1.02 (0.98, 1.06) | 1.01 (0.98, 1.03) |
| p75 vs p25 | 4,430 | 0.34 (-0.73, 1.41) | 0.10 (-0.98, 1.18) | -0.18 (-1.08, 0.72) | -0.25 (-1.12, 0.63) | 1.00 (0.99, 1.01) | 1.00 (0.99, 1.00) |
| **Styrene, MA** |  |  |  |  |  |  |  |
| Q1 (4.17, 87.61) | 1,108 | 0.00 (referent) | 0.00 (referent) | 0.00 (referent) | 0.00 (referent) | 1.00 (referent) | 1.00 (referent) |
| Q2 (87.62, 121.31) | 1,107 | -0.96 (-2.61, 0.68) | -0.75 (-2.38, 0.89) | -1.47 (-2.62, -0.32) | -1.32 (-2.35, -0.29) | 0.99 (0.96, 1.02) | 1.00 (0.98, 1.03) |
| Q3 (121.32, 163.12) | 1,107 | 0.84 (-0.66, 2.34) | 0.67 (-0.76, 2.10) | 0.23 (-1.12, 1.58) | 0.14 (-1.14, 1.42) | 1.00 (0.97, 1.03) | 1.00 (0.98, 1.03) |
| Q4 (163.13, 27778) | 1,108 | 1.28 (-0.50, 3.06) | 1.12 (-0.63, 2.87) | 0.36 (-1.01, 1.73) | 0.45 (-0.81, 1.70) | 1.05 (1.01, 1.08) | 1.03 (1.01, 1.06) |
| p75 vs p25 | 4,430 | 0.85 (-0.33, 2.02) | 0.56 (-0.62, 1.74) | 0.51 (-0.29, 1.32) | 0.48 (-0.25, 1.20) | 0.98 (0.95, 1.02) | 1.00 (0.97, 1.02) |

Metabolite long names: CEMA, N-Acetyl-S-(2-carboxyethyl)-L-cysteine; 3HPMA, N-Acetyl-S-(3-hydroxypropyl)-L-cysteine; PMA, N-Acetyl-S-(phenyl)-L-cysteine; MU, *t,t*-Muconic Acid; DHBMA, N-Acetyl-S-(3,4-dihydroxybutyl)-L-cysteine; MHBMA3, N-Acetyl-S-(4-hydroxy-2-butenyl)-L-cysteine; HPMMA, N-Acetyl-S-(3-hydroxypropyl-1-methyl)-L-cysteine; PGA, Phenylglyoxylic acid; MA, Mandelic acid

Likewise, for 1,3-butadiene, the urinary metabolite DHBMA was associated with higher SBP and a higher prevalence of hypertension. When comparing the highest to lowest quartile of DHBMA, the MD in SBP was 2.46 (1.01, 3.92) mmHg and the PR of hypertension was 1.05 (1.01, 1.09). Estimates for SBP and hypertension were also statistically significant per log IQR-change and, although attenuated, generally remained significant after further adjustment for CVD risk factors. The linear association apparent in the quartile models was also observed with the restricted quadratic spline models, with clear positive dose-response relationships with SBP and PR of hypertension observed for DHBMA above 252 ng/mg creatinine (Figures 1 and 2). For the other 1,3-butadiene metabolite, MHBMA3, results were less consistent, but there was an association with hypertension per log IQR of MHBMA3 (PR [95%CI]: 1.01 [1.00, 1.02]).

For the other priority metabolites, results were less consistent. The benzene metabolite, MU, was associated with higher SBP per log IQR of MU (MD [95%CI]: 0.76 [0.05, 1.48] mmHg), but not with higher prevalence of hypertension. When comparing the highest quartile to the lowest quartile, the crotonaldehyde metabolite, HPMMA, and the styrene metabolite, MA, were associated with higher prevalence of hypertension (PR [95%CI]: 1.06 [1.02, 1.10], and 1.05 [1.01, 1.08]), respectively. Estimates for HPMMA and MA and SBP trended positive but were not statistically significant. Additionally, we found no significant results for any of the priority VOC metabolites and DBP (Table 3 and Figure S3). Results for the other urinary VOC metabolites available in NHANES (i.e., parent compounds: acrylamide, acrylonitrile, 1-bromopropane, carbon disulfide, cyanide, n,n-dimethylformamide, isoprene, propylene oxide, toluene, and xylene) are reported in Table S3.

In Wald tests for interaction, we found significant interactions between age and all priority VOC metabolites except for the benzene metabolite PMA (Table S4). In models stratified by age groups for priority VOCs, the associations with SBP were stronger for the age subgroup 40-64 for metabolites of benzene (MU), 1,3-butadiene (DHBMA), and styrene (PGA). For hypertension, associations were stronger for the age group 40-64 and ≥65 for metabolites of acrolein (CEMA) and crotonaldehyde (HPMMA), the age group 40-64 for the metabolites of benzene (MU) and styrene (PGA), and the age group ≥65 for metabolites of 1,3-butadiene (MHBMA3) and styrene (MA). We found significant interactions between sex and metabolites of acrolein, 1,3-butadiene, and crotonaldehyde. By sex, the association with SBP was stronger in females for metabolites of acrolein (CEMA, 3HPMA), 1,3-butadiene (DHBMA) and crotonaldehyde (HPMMA). We found significant interactions between race and ethnicity for all VOC metabolites except for the benzene metabolite PMA and the styrene metabolite PGA. The association with SBP was stronger for Mexican Americans for 1,3-butadiene (DHBMA), for Non-Hispanic Asians for acrolein (CEMA), and for Non-Hispanic Blacks for styrene (MA). The association with hypertension was stronger for Non-Hispanic Whites for metabolites of acrolein (3HPMA), benzene (MU), crotonaldehyde (HPMMA) and styrene (MA). Patterns for other VOCs were inconsistent.

In sensitivity analyses, we found that effect sizes of associations with underlying BP were consistent with the main analyses of original, measured BP (data not shown). Similarly, the use of Stage 2 or physician diagnosed definitions of hypertension did not significantly change effect sizes; however, use of stage 1 hypertension outcome attenuated associations, except for the styrene metabolite (MA) (Figure S4).

Urinary VOC metabolites originating from the same parent compound were highly correlated except for 1,3-butadiene metabolites, where MHBMA3 was more highly correlated with crotonaldehyde metabolite, HPMMA (Figure S2). We found no significant association between the VOC metabolite mixture and SBP (Figure S5). However, the VOC mixture and overall risk of hypertension was significant and positive. Conditional posterior inclusion probabilities were higher for acrolein metabolite, CEMA, and 1,3-butadiene metabolite, DHBMA, for SBP and CEMA for hypertension (Table S5).

## DISCUSSION

In this nationally representative study of non-smoking adults, we found that the acrolein metabolite, CEMA, and the 1,3-butadiene metabolite, DHBMA, were consistently and significantly associated with higher SBP and higher prevalence of hypertension in quartile, log IQR, and restricted spline models. The other priority VOC metabolites were less consistent across BP outcomes. Estimates were attenuated after adjustment for cardiovascular risk factors. Exploratory hierarchical BKMR analyses supported acrolein and 1,3-butadiene as important contributors to SBP and hypertension risk and the overall mixture of VOC metabolites (excluding benzene) was associated with hypertension.

Elevated BP is an important risk factor for the development of CVD and is associated with the highest population attributable fraction for CVD deaths in the U.S. (40.6%).^36^ Systolic hypertension is usually the result of arterial thickening and stiffening caused either by atherosclerosis, medial degradation, or impaired endothelial-mediated vasodilation,^37^ processes that have been shown to be sensitive to VOCs in experimental studies.^14, 38^ Taken together, these observations suggest that exposure to VOCs, particularly acrolein and 1,3-butadiene, may be significant contributors to the development of CVD, in the general non-smoking population.

Among NHANES 2011-2018 participants the acrolein metabolite, CEMA, was associated with higher SBP and prevalence of hypertension and the 3HPMA metabolite was associated with higher prevalence of hypertension. Acrolein is a pesticide, combustion byproduct, precursor in the production of other chemicals, and byproduct of lipid peroxidation, glycation, and amino acid oxidation; 3HPMA and CEMA account for about 30% of the dose from exposure.^39^ Epidemiological evidence shows acrolein metabolites are associated with SBP among nonsmoking adults, including in a cohort of 308 adults in Louisville, KY of mean age 51.7 years (MD [95%CI] CEMA: 1.2 [-0.3, 3.3] mmHg and 3HPMA: 0.98 [0.04, 1.9] mmHg)^14^ and a cohort of 778 Black participants in Jackson, MS of mean age 51.3 years (CEMA: 1.6 [0.4, 2.7] mmHg and 3HPMA: 0.8 [0.01, 1.6] mmHg).^40^ However, associations between acrolein metabolites and hypertension were null among participants in Jackson, MS. A 2022 analysis of ambient VOC exposure in the Sister Study (n=47,467) found that ambient acrolein exposure was associated with a greater risk of hypertension, particularly among never smoking women 35-74 years of age.^41^ Likewise, in a nested case-cohort study of non-smoking individuals in northeastern Iran (n=1,198), there was an increased hazard ratio of ischemic heart disease mortality associated with metabolites of acrolein (CEMA, 2.11 (1.31, 3.40) and 3HPMA, 2.99 (1.60, 5.59)) when comparing the highest tertile to the lowest.^18^

In this study, we also found the 1,3-butadiene metabolite, DHBMA, was positively associated with higher SBP and greater risk of hypertension, and MHBMA3 was associated with higher prevalence of hypertension. Most human exposure from 1,3-butadiene occurs from incomplete combustion of fossil fuels and burning of biomass.^42^ DHBMA is the most abundant metabolite in urine due to its natural background, which may be caused by endogenous sources.^43, 44^ It is unknown whether high background levels of DHBMA are due to 1,3-butadiene exposures or endogenous sources like lipid peroxidation. Additionally, crotonaldehyde is produced endogenously from the metabolism of 1,3-butadiene and may contribute to higher levels of DHBMA.^45–47^ Although MHBMA3 is a sensitive marker of 1,3-butadiene exposure, DHBMA is not a sensitive marker,^48^ and does not decrease with smoking cessation, compared to MHBMA3.^49^ Therefore, DHBMA may be a relevant biomarker for CVD risk,^50^ but may not be indicative of 1,3-butadiene exposure. In the Sister Study (n=47,467), ambient 1,3-butadiene at the census tract level was associated with 1.05 (95% CI 1.00, 1.10) greater risk of hypertension.^41^ Additionally, there was a higher ischemic heart disease mortality hazard ratio associated with 1,3-butadiene [DHBMA, 2.49 (1.50, 4.15)] in a recent study of non-smoking northeastern Iranians (n=1,198).^18^ However, associations were null among participants in the Louisville, KY and Jackson, MS studies.^14, 40^ More studies of low-level 1,3-butadiene exposures are required to better assess the sensitivity of DHBMA as an exposure biomarker.

The other priority metabolites were associated with either higher SBP or higher prevalence of hypertension, but not both. The benzene metabolite, MU (but not PMA), was associated with higher SBP. This may be due to smaller sample size compared with the other VOC metabolites. Limited, consistent evidence exists of urinary benzene metabolites and ambient benzene and cardiovascular outcomes due to ongoing analytical issues.^51, 52^ In the Sister Study, ambient benzene was significantly associated with a 1.06 (95% CI 1.01, 1.10) greater risk of hypertension among never smokers.^41^ The crotonaldehyde metabolite, HPMMA, was associated with higher prevalence of hypertension. Among nonsmoking Black participants in Jackson, MS, HPMMA was associated with higher SBP (MD: 1.2 [95%CI: 0.08, 1.8] mm Hg),^40^ but associations with hypertension were null. Similarly, results for HPMMA and BP were null in the Louisville, KY cohort.^14^ The styrene metabolite, MA (but not PGA), was associated with higher prevalence of hypertension. Consistent with our results, the Consortium of Safe Labor showed that ambient styrene was associated with greater odds of hypertension and higher SBP and DBP among normotensive women.^53, 54^ Additionally, two longitudinal studies of occupational co-exposure to 1,3-butadiene and styrene have shown increased standardized mortality ratios for arteriosclerotic heart disease in non-White populations;^55–57^ there was no data on only styrene exposure. However, in the recent case-cohort study in Iran, there was an increased risk of ischemic heart disease mortality associated with both metabolites of styrene [MA, 1.94 (1.18, 3.20) and PGA, 1.54 (1.00, 2.37)].^18^

### Limitations & Future Direction

This large study of VOC metabolites and blood pressure in a representative population of U.S. adults included 4,430 nonsmoking participants and evaluated 35 different VOC metabolites in urine. To minimize error from multiple testing, we prioritized nine VOC metabolites from five parent VOCs, acrolein, benzene, 1,3-butadiene, crotonaldehyde, and styrene, based on prior evidence. All VOC metabolites with >50% observations above LOD available in NHANES, however, were explored in secondary analyses. Previous studies have measured VOCs in air and grouped them by structure, functional group, or as a measure of total VOCs; here, we used stable urinary metabolites of VOC exposure, a robust measure of short-term exposure. The existing literature has been limited by the sample size of non-smokers with measured VOC metabolites. By combining 4 cycles of NHANES we increased the non-smoking sample size resulting in the largest study of VOC metabolites and BP among the general, non-smoking US population. Because metabolites from the same parent VOCs are highly correlated, we used hierarchical BKMR to confirm single-pollutant models after taking correlated VOC co-exposures into account, as well as determined the effects of VOC mixtures on BP and hypertension. These mixture analyses were only exploratory as a weighting scheme for complex survey designs has not yet been developed for BKMR.

Other limitations include the cross-sectional design and potential for exposure misclassification. Data from other cohorts designed to study cardiovascular health, like the Multi-Ethnic Study of Atherosclerosis, may help elucidate differences in cardiovascular risk among VOC exposure groups, particularly with a longitudinal design to evaluate changes in BP levels and hypertension risk over time.

## PERSPECTIVES

This study in NHANES found that metabolites of acrolein and 1,3-butadiene were associated with higher SBP and risk of hypertension, in the non-smoking, general US population. Additional research is needed to characterize the contribution of non-smoking related environmental VOC exposures to cardiovascular risk and to develop policy and interventions to curb preventable exposure to VOCs and further reduce the risk of CVD.

## NOVELTY AND RELEVANCE

### What is New?

- This is the largest biomonitoring study of VOC exposure and elevated blood pressure in the general, non-smoking U.S. population.
- VOCs present in the environment and/or endogenous VOCs are associated with higher systolic BP and greater prevalence of hypertension.

### What is Relevant?

- VOC exposure, independent of cigarette smoking, may contribute to the growing prevalence of cardiovascular diseases

### Clinical/Pathophysiological Implications

- Reduction of VOC exposure may improve cardiovascular health.

## Data Availability

All data is available for download from the National Center for Health Statistics (NCHS), Centers for Disease Control and Prevention (CDC) website.

https://wwwn.cdc.gov/nchs/nhanes/

## NON-STANDARD ABBREVIATIONS AND ACRONYMS

3HPMA: N-Acetyl-S-(3-hydroxypropyl)-L-cysteine
BMI: Body mass index
BKMR: Bayesian Kernel Machine Regression
BP: Blood pressure
CEMA: N-Acetyl-S-(2-carboxyethyl)-L-cysteine
CI: Confidence intervals
CVD: Cardiovascular disease
DBP: Diastolic blood pressure
DHBMA: N-Acetyl-S-(3,4-dihydroxybutyl)-L-cysteine
eGFR: Estimated glomerular filtration
HDL: High density lipoprotein cholesterol
HPMMA: N-Acetyl-S-(3-hydroxypropyl-1-methyl)-L-cysteine
MA: Mandelic acid
MHBMA3: N-Acetyl-S-(4-hydroxy-2-butenyl)-L-cysteine
MU: t,t-Muconic Acid
NHANES: National Health and Nutrition Examination Survey
PGA: Phenylglyoxylic acid
PIPs: Posterior inclusion probabilities PMA – N-Acetyl-S-(phenyl)-L-cysteine
SBP: Systolic blood pressure
VOCs: Volatile organic compounds

## ACKNOWLEDGEMENTS

Data and materials produced by Federal agencies are in the public domain and may be reproduced without permission. However, the authors would like to thank participants in the National Health and Nutrition Examination Surveys (NHANES).

## SUPPLEMENTAL MATERIAL

Tables S1-S5

Figures S1-S5

